# The effect of conflict on medical facilities in Mariupol, Ukraine: a quasi-experimental study

**DOI:** 10.1101/2023.08.01.23293508

**Authors:** Danielle N. Poole, Daniel Andersen, Nathaniel A. Raymond, Jack Parham, Caitlin Howarth, Oona A. Hathaway, Kaveh Khoshnood, Yale Humanitarian Research Lab

**Author notes:** Corresponding author Danielle N. Poole Yale Humanitarian Research Lab Epidemiology of Microbial Diseases Department Yale School of Public Health 60 College St New Haven CT, USA 06510 E.

## Abstract

Medical facilities are civilian objects specially protected by international humanitarian law. Despite the need for systematic documentation of the effects of war on medical facilities for judiciary accountability, current methods for surveilling damage to protected civilian objects during ongoing armed conflict are insufficient. Satellite imagery damage assessment confers significant possibilities for investigating patterns of war. We leveraged commercially and publicly available satellite imagery and geolocated facility data to conduct a pre-post quasi-experimental study of damage to medical infrastructure in Mariupol, Ukraine as a result of Russia’s invasion. We found that 77% of medical facilities in Mariupol sustained damage during Russia’s siege lasting from February 24 - May 20, 2022. Facility size was not associated with damage, suggesting that attacks on medical facilities are not a residual of physical infrastructure characteristics. This is the first geographically comprehensive pre-post study of the effects of an ongoing conflict on specially protected medical infrastructure.

---

Medical facilities are civilian objects that are specially protected by international humanitarian law. Moreover, medical personnel and the wounded and sick must be respected and protected and therefore cannot be intentionally targeted. These protections are enshrined in the 1949 Geneva Conventions^1–3^ and the two additional protocols to the Conventions.^4, 5^ Serious violations of these protections, including attacks on medical facilities, can constitute war crimes. And yet, violence against medical facilities has been employed in armed conflicts around the world for decades^6–9^ and appears to be on the rise.^10^

Surveillance of attacks on medical facilities is a widespread mechanism for accountability and deterrence.^11^ Resolutions from the World Health Assembly, the United Nations (UN) Security Council, and the UN General Assembly have reiterated the need to compile data on these violations to protect health in conflict.^12–15^ With the mandate to provide leadership on the collection and dissemination of data about attacks on medical facilities, the World Health Organization established the Surveillance System for Attacks on Health Care in 2015. Additional platforms to document attacks on medical facilities are hosted by a limited group of international organizations.

Despite interest in the documentation of attacks on medical facilities,^8^ coordinated and systematic data collection efforts have lagged. Current mechanisms for surveilling attacks on protected civilian objects are insufficient with sensitivity as low as 1%.^16, 17^ Moreover, the absence of standardized incident classification and verification across data sources, challenges to responsibly sharing location information in conflict settings, and privileging of particular forms and locations of attacks limit validity.^18, 19^ Evidence reported in the peer-reviewed literature is similarly weak, with only 11% of studies collecting quantitative data. These weaknesses have produced a restricted, non-representative evidence base^20^ with a particular dearth of quantitative data.^8^ The absence of estimated of the impact of conflict on protected medical facilities makes it difficult to identify trends,^21^ weakens efforts to prevent future attacks,^11, 18^ and limits the potential of accountability mechanisms.

Unless attacks are systematically documented, there will continue to be an important gap in knowledge regarding the extent of damage to medical facilities during armed conflict.^21^ This gap, if left unaddressed, will have consequences for the provision of medical assistance during ongoing conflict and efforts to reconstruct health systems after armed conflict. Moreover, this gap will undermine efforts to hold alleged perpetrators accountable. Satellite imagery damage classification is often the only pathway available for empirically detecting and further investigating the effects of conflict on medical facilities.^22, 23^ Such remote assessment is increasingly central to the collection of evidence of alleged war crimes and human rights violations,^24, 25^ including as evidence in cases before the International Criminal Tribunals for the former Yugoslavia, the International Criminal Court, and the International Court of Justice.^26–28^ Today, near real-time, georeferenced damage classification can be performed in nonpermissive conflict environments using publicly available satellite imagery.^29^

We leverage commercially and publicly available satellite imagery and medical infrastructure databases to determine the prevalence of attacks on medical facilities during the siege of Mariupol in a pre-post quasi-experimental study. Previous investigations have reported case-based findings of attacks on medical facilities. We overcome this limitation by generating a comprehensive, georeferenced dataset of all medical facilities within a geographic area of interest, the city of Mariupol, during the siege lasting from February 24 - May 20, 2022. Geographic locations of medical infrastructure were acquired and cross-corroborated in OpenStreetMap (OSM), Google Maps, Wikimapia, and a locally maintained dataset recently published by the Ukrainian Healthcare Center (UHC).^30^ Facility size was calculated as overall volume, area, and maximum height using the building footprint and floor polygons obtained from OSM. We then used an established methodology^31^ to assess structural damage to all medical facilities within the city of Mariupol during the study period. Lastly, we spatially matched all incidents of damage to events in the Armed Conflict Location and Event Data (ACLED) Project^32^ to identify the associated actor and event type.

## Medical facility damage census

A total of 73 medical facilities were geolocated and cross-corroborated within the city of Mariupol. Over three-quarters (77%; 95% CI: 65-86%) of medical facilities sustained damage during the study period. **Figure 1** presents an example of highest commercially available resolution satellite imagery remote detection of damage to a medical facility.^33^

**Figure 1.**
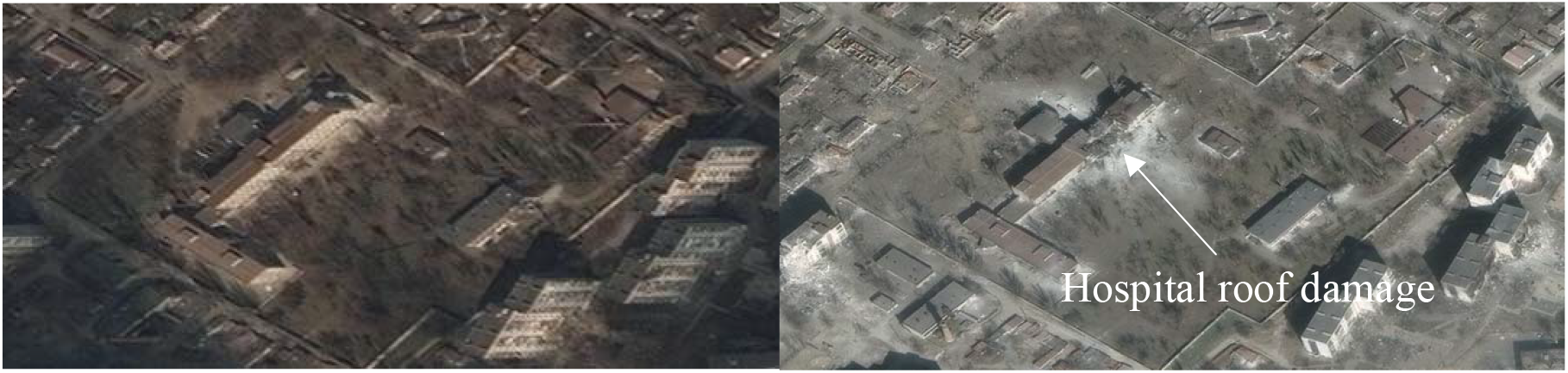
Damage to the Mariupol Maternity Hospital consistent with direct impact from bombardment. ©2022 Maxar Mariupol Maternity Hospital on March 18, 2022 (left) and March 29, 2022 (right).

## Facility characteristics

Medical facility characteristics, derived from OSM, are presented in **Table 1**. The median building was 10,819 cubic meters in volume, with a footprint area of 1,014 meters squared, and a median height of three floors (or, approximately 12 meters).

**Table 1.**
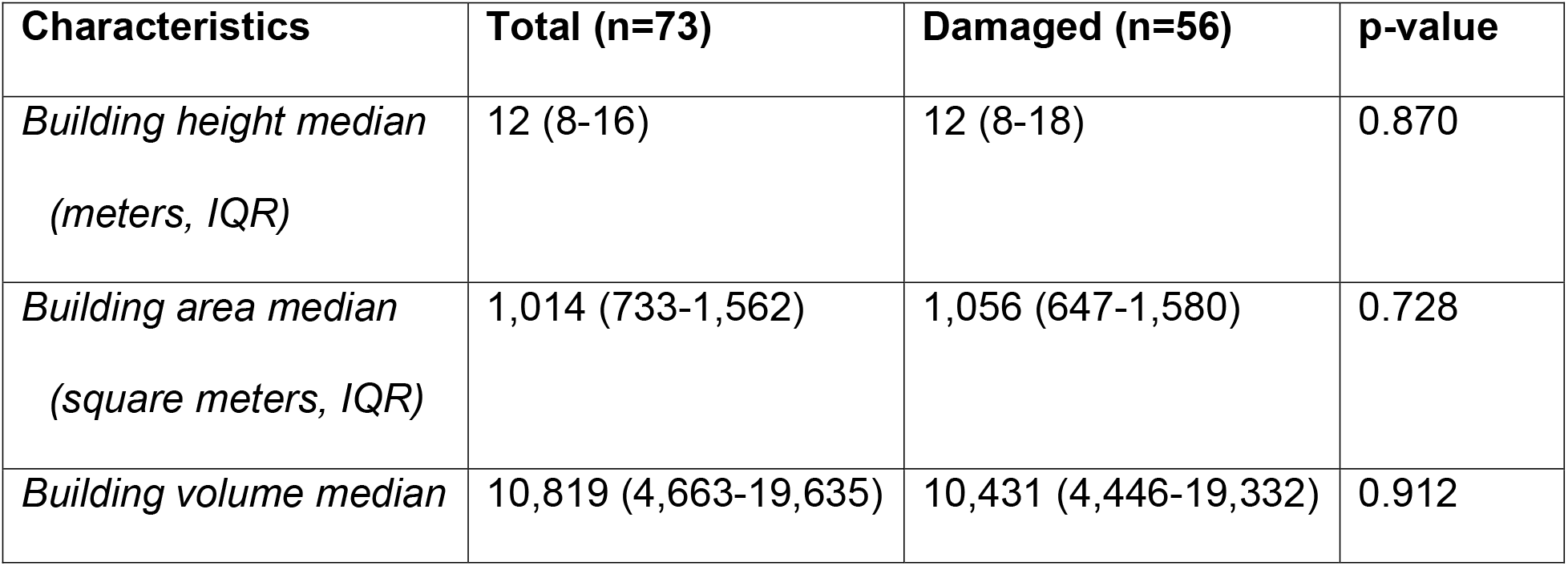
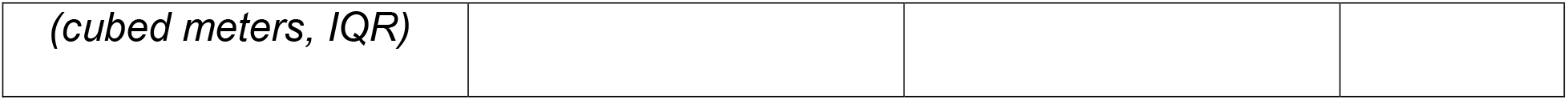
Medical facility characteristics and associated damage.

| **Characteristics** | **Total (n=73)** | **Damaged (n=56)** | **p-value** |
| --- | --- | --- | --- |
| *Building height median*<br>*(meters, IQR)* | 12 (8-16) | 12 (8-18) | 0.870 |
| *Building area median*<br>*(square meters, IQR)* | 1,014 (733-1,562) | 1,056 (647-1,580) | 0.728 |
| *Building volume median* | 10,819 (4,663-19,635) | 10,431 (4,446-19,332) | 0.912 |
| (cubed meters, IQR) |  |  |  |

There was no significant difference in the building size, including the overall volume, height, and footprint, of medical facilities that sustained damage and those that did not. The lack of difference in the size of facilities that sustained damage is suggestive of intentional targeting of medical facilities, which would constitute a war crime, as random saturation of artillery fire or unintentional damage would be expected to hit larger facilities with a larger footprint at a higher frequency.

## Event type and actor

Of the 56 damaged medical facilities, 46 (82%) were geospatially matched with ACLED conflict data^32^ describing the event type and actor. All of these geospatially matched events were attributed to the Armed Forces of the Russian Federation, comprising 31 incidents of an “Air/drone strike” and 19 incidents of a “Shelling/artillery/missile attack.”

## Discussion

We demonstrate the feasibility of generating systematic and reproducible estimates of the effects of conflict on medical facilities using commercially and publicly available data sources. We found a preponderance of evidence of damage to medical facilities, which are specially protected civilian objects, in the city of Mariupol. Each incident of an attack on a medical facility may represent an individual and distinct criminal act under international humanitarian law.^1–5^ The percentage of damaged medical facilities identified in this study is consistent with previous reports of destruction to medical facilities and infrastructure in Mariupol, ranging from 80-90%;^30, 34^ however, variability in the sampling and damage assessment methods precludes direct comparison of these estimates.

This study identified incidents occurring when Mariupol was under siege by Russia.^36^ Geospatially matched ACLED event and attribution data identified the Armed Forces of the Russian Federation as the actor associated with the vast majority of aerial strikes corresponding to the damage observed in this study.

Eighty-four percent of attacks on medical facilities in Ukraine are reported to have been performed with heavy weaponry, typically trained on the largest structures.^35^ However, our findings demonstrate that damage was not associated with measures of building size, suggesting that larger buildings did not have a higher likelihood of being damaged as an artifact of structural features. Therefore, is not a determinant of attacks on medical facilities.

Medical facility locations are increasingly maintained in online databases with information such as building latitude and longitude, footprint polygons, and imagery of signage.^37, 38^ The locations of the medical facilities included in this study are available to the public, including to the Armed Forces of the Russian Federation, in a minimum of two databases. Together, these findings are consistent with reports of targeted attacks on medical facilities, including the failure to take precautions with respect to specially protected civilian objects, during Russia’s invasion of Ukraine.^39^

### Strengths

A major strength of our study is the pre-post quasi-experimental study design used to estimate the effects of Russia’s siege on medical facility infrastructure by applying reproducible methods to publicly available data sources. The resulting scientifically rigorous findings are expected to foster a continuum of knowledge, evidence, and practice that can support war crime accountability.^21^ Moreover, the analysis of publicly available data for the generation of evidence could be made available to all actors—not just the prosecution, but defense as well—and in a variety of accountability settings, from international to domestic and from courtrooms to diplomatic settings.

### Limitations

This study has several limitations. First, damage assessments conducted primarily using satellite imagery are limited to features visible to analysts and may not always show damage, even extensive damage, to a structure’s sides and interior, and may be obscured by the presence of cloud cover or other environmental interference. Thus, this study provides a conservative estimate of damage. Combining this data with ground-level assessments and additional sensors, including medical facility functionality indicators, would more accurately capture the scope of damage. Nevertheless, cyber forensics capacity, including the analysis of satellite imagery, represents an important source of evidence for national and international criminal justice.^24, 40^ Second, war crimes prosecution requires not only identification of the individual perpetrator responsible, but also a showing of that individual’s criminal intent, both of which are beyond the scope of this study. Third, and related, this study does not assess whether the identified medical facilities may have been used for non-medical military purposes, thus becoming a lawful military object, or whether there may have been adjacent buildings that constituted lawful military objects that may have been the true targets of the attacks. Yet, even if Russia’s armed forces had military objectives in conducting the attacks, the special protected status of medical facilities should have resulted in precautionary care. In addition, the international humanitarian law principle of proportionality – which prohibits attacks against military objects that are expected to cause incidental harm to civilians or civilian objects that would be excessive in relation to the concrete and direct military advantage anticipated – likely would have rendered the attacks unlawful.^39^

### A way forward for war crimes accountability

Even recognizing these limitations, this study offers an important way forward for accountability for violations of international humanitarian law—and thus, it is hoped, for deterring future violations. A key difficulty in war crimes prosecutions has long been evidence collection. During ongoing conflict, evidence can be difficult, if not impossible, to gather—and doing so can pose serious dangers to investigators. Moreover, given limited and sporadic access to the site of violations, it can often be challenging to pinpoint when a violation took place: information that is necessary to building evidence that can be used in a criminal prosecution. This study demonstrates that comprehensive, reproducible evidence of potential violations of international humanitarian law can be gathered using publicly available satellite imagery together with other public data sources. In doing so, it offers an important solution to longstanding problems in providing accountability for violations of the law.

It is important to recognize that evidence of potential violations of international humanitarian law is a necessary, but not sufficient, step towards war crime accountability. Most important, cases must be brought against perpetrators – either in international courts, such as the International Criminal Court, or in domestic courts.^41^ Moreover, for such evidence to be used effectively, courts and those practicing before them must develop expertise in and procedures for gathering, assessing, and preserving these new forms of evidence. For example, integration of methods demonstrating systematic damage to protected civilian objects, such as medical facilities, should be incorporated into the International Criminal Court Rules of Procedure and Evidence to ensure the generation and preservation of admissible evidence.^42^

This study is meant to demonstrate a technique that can be used in a variety of contexts. Future research should seek to expand these methods to document other legal violations, including war crimes beyond the targeting of medical facilities and violations of international human rights law, accountability for which often suffers from the same evidentiary hurdles.

### Conclusions

To our knowledge, this evidence of the widespread destruction of medical facilities is the first pre-post quasi-experimental study of the effects of war on medical infrastructure. As such, the reproducible methods, incorporating publicly available data, have the potential to make substantial contributions to the international justice system by providing real-time estimates of damage to specially protected civilian objects. By combining the strengths of satellite imagery analysis in nonpermissive environments with publicly available data sources of medical facility infrastructure, this study demonstrates the power of data amalgamation for creating outputs that were, until recently, inaccessible. However, because the impact of this work is contingent upon its use in international justice mechanisms, this study also represents a call for action to clarify the forensic standards for remote documentation of war crimes and expand expertise in collecting and assessing such evidence.

## List of abbreviations

95% CI: 95% Confidence Interval
ACLED: Armed Conflict Location and Event Data Project
IQR: Interquartile Range
OSM: OpenStreetMap
UHC: Ukrainian Healthcare Center
UN: United Nations

## Data Availability

A de-identified dataset generated during the current study is available from the corresponding author on reasonable request.

## Acknowledgements

This study is a product of the members of the Yale Humanitarian Research Lab who are leading the advancement of war crime documentation. This independent study builds upon ongoing work supported by the US State Department and Conflict Observatory consortium. This study does not necessarily reflect the views of the United States Government.

## Author contributions

DNP designed the study, analyzed and interpreted the data, and wrote the manuscript. DA and JP produced and analyzed the data. NAR and KK contributed to the conceptual design of the study. CH and OH interpreted the data. Anonymous members of the Yale HRL contributed to the conceptual design of the study and acquired, analyzed, and interpreted the data. All authors provided critical review and assisted in writing the manuscript.

## Materials & Correspondence

Correspondence and material requests should be addressed to Dr. Danielle N. Poole.

## Methods

This study includes a census of damage to medical facilities within the city of Mariupol, located in the Donetsk Oblast of Ukraine during the siege beginning on February 24 and lasting until May 20, 2022, as part of Russia’s invasion of Ukraine. The city of Mariupol has an area of 166 km. The population of Mariupol has decreased five-fold since the invasion, from 425,000 to 85,000 inhabitants.^30^ Prior to the start of the war in 2022, the city had an extensive network of medical care facilities including five city hospitals, one regional hospital, a network of six primary care centers with multiple service points, specialized facilities providing maternal and pediatric care, and private medical facilities.^30^

### Study design

This observational, pre-post quasi-experimental study identifies the effects of Russia’s invasion on medical facilities within Mariupol.

### Data sources

Publicly available georeferenced medical facility data and satellite imagery were combined to generate an empirical census of damage to medical facilities in the study setting.

#### Administrative boundaries

Shapefiles storing the attribute information of geographic features, including administrative boundaries of Mariupol, were obtained from Natural Earth.^43^

#### Exclusion and inclusion criteria

Comprehensive, georeferenced sources of medical facility data vary in quality and completeness.^44^ Moreover, these limitations are exacerbated by conflict, and compounded by limited data sharing of medical facilities located in war zones. We used medical facility location data from multiple, publicly available databases including (1) OpenStreetMap (OSM), (2) data from Google Maps, (3) Wikimapia, and (4) published medical facility datasets from the Ukrainian Healthcare Center (UHC) to generate a cross-corroborated dataset. Of note, past examples of geolocated medical facilities have relied on a single data source, although such databases are known to have heterogenous content and quality. Cross-corroboration of facility type and location increases the validity of our findings.

First, medical facility polygon and latitude and longitude point layers were extracted from the OSM database using facility-type attribute. The OSM dataset is a collaborative project designed to create a free and editable geospatial database of the whole world. OSM is one of the most successful examples of a volunteered geographic information project built by a large user community that employs aerial imagery, GPS devices, and low-tech field maps to verify that OSM is accurate and up to date.

Facilities were included in the study sample if they were categorized as hospitals, clinics, or facilities providing urgent or emergency medical care with a database entry subtype indicating they were a hospital or clinic. This dataset was merged with the geocoordinates of medical facilities documented by UHC.^30^ Facilities included in both OSM and UHC databases were considered verified by multiple sources. Facilities included in only OSM or UHC databases were further verified in Google Maps and Wikimapia. Facilities identified in only one database were excluded. The resulting dataset included 73 facilities identified in two or more databases.

The underlying sources of the geolocated entities within Google Maps included publicly available data, licensed third-party data and data contributed by users ^45^ Publicly available and third-party data may be associated with dataset-specific metadata that describe their accuracy and completeness, and, as with OSM, users of Google Maps can flag and report potential errors.

#### Facility characteristics

Building footprint data were derived from OpenStreetMap.^46^ Area was calculated by summing the area of the polygons for each medical facility. Building height was calculated by multiplying the highest floor by 3.9 meters.^47^ Google Maps and Yandex were used to determine the number of floors for polygons missing this information. Volume was calculated for each polygon and summed to generate the medical facility volume.

#### Damage classification

Every facility identified and verified through open source documentation was assessed via satellite imagery analysis to establish a baseline structural assessment. The very high resolution imagery used to support this investigation was unclassified, commercially available imagery captured by Maxar Technologies, Planet Labs, and other commercial satellite imagery suppliers provided by the US Department of State to the Humanitarian Research Lab at Yale University School of Public Health. Latitude/longitude was recorded for each medical facility.

Multi-temporal change detection was conducted by a minimum of two geospatial analysts. Indicators of damage to infrastructure include changes in feature coloration, texture, and pattern as seen from above. Spatial resolution between 38 and 50 cm and temporal resolution in this imagery allowed analysts to assess changes in infrastructure and the natural environment, both of which may visibly reveal damage caused by heavy weapons and some small arms commonly utilized in armed conflict.

Damage visible in satellite imagery was used to classify medical facilities as sustaining: 1) no visible damage; 2) possible damage; and 3) damage. These categories were defined by the damage assessment scale developed by the International Working Group on Satellite-based Emergency Mapping (IWG-SEM).^31^ No distinction was made between minor and major damage classification due to variability in imagery resolution.

**Table 1.**
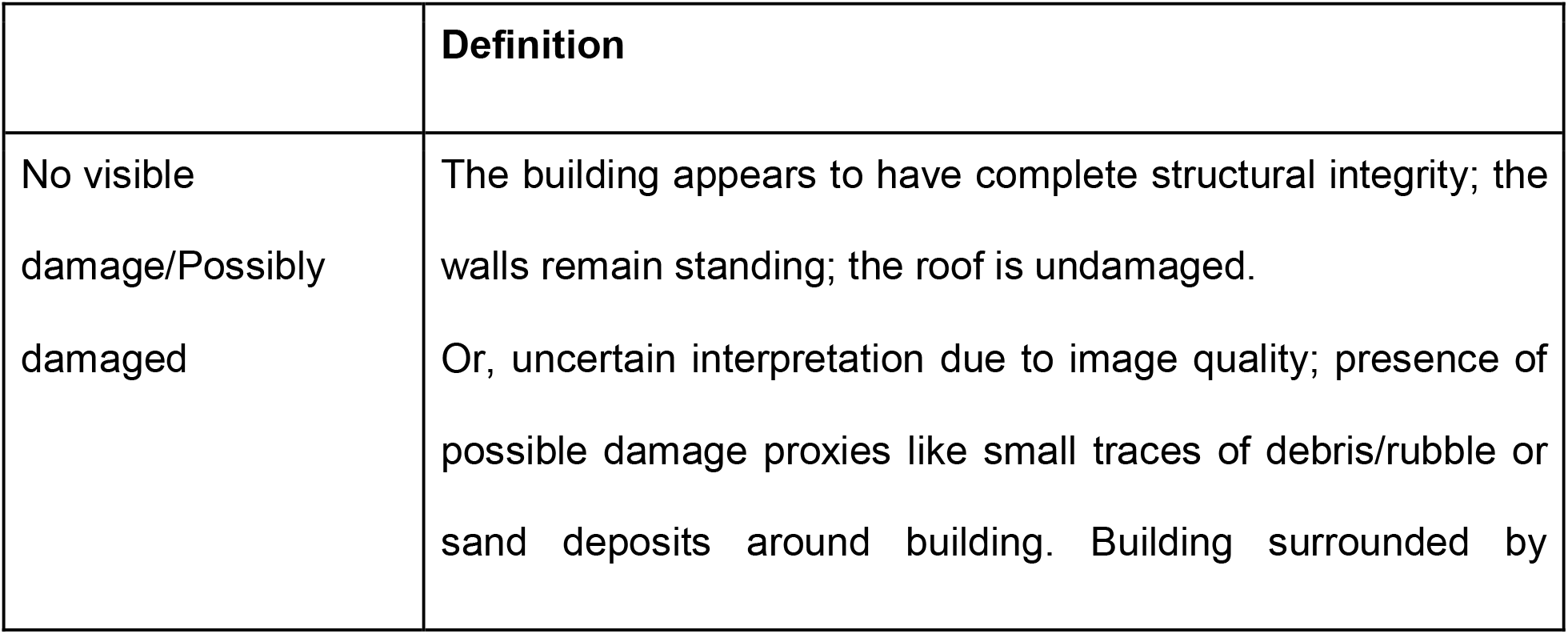
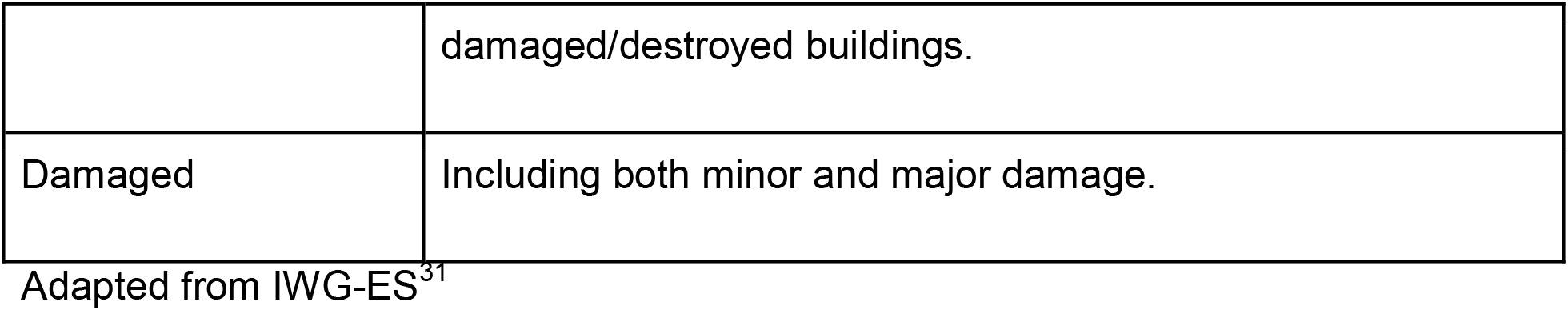
Damage classification.

|  | Definition |
| --- | --- |
| No visible damage/Possibly damaged | The building appears to have complete structural integrity; the walls remain standing; the roof is undamaged.<br>Or, uncertain interpretation due to image quality; presence of possible damage proxies like small traces of debris/rubble or sand deposits around building. Building surrounded by |
|  | damaged/destroyed buildings. |
| Damaged | Including both minor and major damage. |

Adapted from IWG-ES<sup>31</sup>

Damage assessments were repeated on all available imagery throughout the study period. Agreement on damage classification status during the study period was achieved from a minimum of two analysts.

#### Event type and actor

The primary incidence of damage to a medical facility was geographically matched to publicly available event data recorded in the Armed Conflict and Location and Event Data Project (ACLED). The ACLED Project collects, analyzes, and maps the incidence and characteristics of a range of violent events, from inter-state battles to riots and other forms of civil unrest.^32^ We used ACLED date, location (latitude/longitude), type, and actor variables for conflict events in Ukraine from February 24 – May 20, 2022.

### Statistical analysis

This study aimed to conduct a damage census, including all medical facilities in the study setting. The proportion of damaged medical facilities was calculated with Clopper-Pearson 95% CI for binomial proportions. The study variables were analyzed using frequency, percentage, mean, median, and interquartile range (IQR). Logistic regression models assessed associations between categorical damage and building characteristics. Ordinary least squares regression models assessed the relationship between categorical damage and continuous variables. All analyses were conducted in Stata version 17.0.^48^

